# Biomarkers of High Flow Therapy Outcome in COVID-19 pneumonia: a single-center prospective study

**DOI:** 10.1101/2024.07.13.24310359

**Authors:** Toni. Marín, Irene Aldás, Marina Galdeano, Agnes Hernández, Leire Mendiluce, Roxana Chirinos, Carmen Fernández, Adriana Martín, Esther Roca, Cristian Tebé, Roger Paredes, Antoni Rosell

## Abstract

Patients with acute hypoxemic respiratory failure (AHRF) due to COVID-19 undergoing High Flow Therapy (HFT) before intubation presented an increased risk of mortality when intubation was delayed. We designed a prospective study seeking biomarkers for early prediction of HFT failure. An analytical value in blood could be more stable than the ROX index since it will not depend on the vital signs that the patient presents at that moment. We defined HFT failure as the need to scale the treatment to Non-Invasive Positive Pressure Ventilation (NPPV). The needs were respiratory rate >25, oxygen saturation of <90% despite being on flow of 60 l·min-1 and FiO2 1 or levels of PaO2/FiO2 ratio <100mmHg. The all-treatment population included all subjects enrolled in the trial. 139 patients were enrolled after starting HFT. The Pearson chi-squared test was used to compare the main study outcomes. These included the incidence of intubation, the cumulative incidence of mortality at 30 days, the cumulative incidence of mortality at 1 year, and the composite outcome of intubation or death at the end of the trial. Kaplan-Meyer plot was used to illustrate the time to HFT failure. The Cox regression model was used to estimate the hazard ratio for HFT failure for all the parameters. All were measured or collected at baseline. Lower levels of bicarbonate, thrombocytopenia, and higher levels of C-reactive protein (CRP), lactate dehydrogenase (LDH), creatinine, and glucose are early blood biomarkers independently associated with HFT failure.

**SUMMARY AT A GLANCE:** Higher levels of C-reactive protein and lactate dehydrogenase in patients with COVID-19 pneumonia allow us to early detect patients requiring intubation with an apparent good response to high-flow oxygen therapy.

## INRODUCTION

In patients with acute hypoxemic respiratory failure (AHRF) due to COVID-19, High Flow Therapy (HFT) seems to be superior to conventional oxygen in reducing the risk of intubation but not in reducing mortality among patients with AHFR. (1–8) Mechanical ventilation is resource-intensive and is associated with higher need for sedation and immobility, which have been associated with higher rates of complications. There may be benefits from preventing intubation, even in the absence of a significant improvement in mortality, including for patients with ‘do-not-intubate’ (DNI) orders. (9) HFT is generally well tolerated. However, HFT before mechanical ventilation presented an increased risk of mortality when intubation was delayed irrespective of the time-point used to consider early or delayed intubation (8,10). It seems important to define the criteria of the patient population that benefits from treatment with HFT. (11) and even more increase our capacity as clinicians to early detect patients who experience HFT failure. Recently Spanish guidelines updated the recommendations based on the evidence using non-invasive respiratory support strategies in the COVID-19 Patient. For patients with PaO2/FiO2 ratio (P/F) less than 200mmHg HFT should be preferred due to better tolerability and fewer side effects. If P/F less than 150mmHg it’s unclear what respiratory support it’s better, but the patient should be closely monitored. (12)

ROX Index is the most used failure predictor of this therapy in all causes of acute respiratory failure (13) also due to COVID-19 (14–16) An analytical value in blood could be more stable than the ROX index since it will not depend on the vital signs that the patient presents at that moment. COVID-19 Spanish ICU Network published in 2021 a multicenter, observational study from a prospectively collected database of consecutive COVID-19 patients who received HFT on ICU admission. Baseline non-respiratory SOFA score, ROX index, and pH were associated with the need for intubation (17,18). There are studies that have explored markers of disease severity. In general, patients with severe or critical COVID-19 have higher levels of inflammatory markers, such as C-reactive protein (CRP), ferritin, lactate dehydrogenase (LDH), and alanine aminotransferase. (19–22) Even so, these studies do not stratify severity based on the respiratory support required. There are no prospective studies that compare biomarkers to predict specifically outcome of HFT.

In 2021, patients with SARS-Cov2 pneumonia, with severe respiratory failure but without intubation criteria at the time of admission, were admitted to the intermediate respiratory care unit. Following the therapeutic strategy proposed by *Scala and Heunks* (23), early initiation of HFT was prioritized over non-invasive positive pressure ventilation (NPPV). The objective of our study was to find novel prognostic factors and find stable biomarkers capable of early predicting HFT failure defined as the need to escalate to NPPV due to respiratory worsening.

## METHODS

### STUDY DESIGN AND SETTING

We conducted a prospective, single-center cohort study of patients with AHRF due to COVID. It was carried out during the months of February and October 2021. Research Ethics Committee of Germans Trias i Pujol University Hospital approved the study with REF PI-20-372. The study was registered on ClinicalTrials.org with NCT05094661. It was conducted according to the amended Declaration of Helsinki. This report follows the “Strengthening the Reporting of Observational Studies in Epidemiology (STROBE)” guidelines for observational cohort studies.

We included consecutive patients over 18 with AHRF due to COVID-19, using standard oxygen FiO2 0.5-1 and P/F <250. We excluded patients in need of immediate intubation, Glasgow coma scale <15, respiratory acidosis, hypercapnia, need to use vasopressors, impaired swallowing, or tracheotomy. Also, patients with home respiratory therapies such as oxygen therapy, CPAP, or NIV.

We started HFT (model AIRVO 2 or V60 plus) with a humidification level of T34-37ºC with a FiO2 OF 1 and a total flow of 30 l·min^-1^ increasing the flow to achieve a respiratory rate of less than 30. We have decreased FiO2 to the minimum necessary to maintain a SpO2 greater than 96%. We recorded variables at 1 h with complete biochemical arterial blood gas and at 48 h. Also, registration of variables at 4h, 6h, 12h, 24h, 48h, after the start of the treatment.

We defined HFT failure as the need to scale the treatment to NPPV. We have switched to NPPV Minimum of 18 hours per day if persistent respiratory work (FR> 25), SpO2 <92% appears at any time after treatment with HFT despite being on flow of 60 l·min^-1^ and FiO2 1; or with the use of accessory muscles or abdominal breathing, and/or subjective exhaustion or levels of P/F <100mmHg. Therapy is terminated too if the patient presents with respiratory improvement that allows the removal of the support. We made weekly follow-ups until day 30 of the study, day 90, and the year after.

## DATA COLLECTION

Patients’ characteristics were collected prospectively from electronic medical records by physicians and nurses trained in critical care according to a previously standardized consensus protocol. The information regarding personal data was coded, and a non-consecutive alphanumeric code was assigned. Patient confidentiality was protected following the provisions of Law 3/2018 on the Protection of Personal Data and Guarantee of Digital Rights and Regulation (EU) 2016/679 of the European Parliament on Data Protection. Recorded data included demographics [age, gender] comorbidities, Charlson Index, disease chronology [time from onset of symptoms and hospital admission to initiation of respiratory support, ICU length of stay], vital signs, ROX index, and severity scores such as the Sequential Organ Failure Assessment (SOFA) were recorded. We obtained variables at baseline and 48 hours after starting HFT, with complete biochemical analysis including blood count, CPR, LDH, Ferritin, IL-6, troponin, CK. After ICU discharge, patients were followed up for 1-year.

## STATISTICAL ANALYSIS

To estimate the proportion of patients undergoing HFT treatment who fail and end in intubation with a precision of +/-5% and an alpha error of 5% 95% CI, a sample size of 351 patients is required.

The demographic and clinical profile of the enrolled subjects was described using standard statistics according to the type of variable. Subject population was defined as all-treatment; that included all subjects enrolled in the trial. The Pearson chi-squared test was used to compare the main study outcomes. These included the incidence of intubation, the cumulative incidence of mortality at 30 days, the cumulative incidence of mortality at 1 year, and the composite outcome of intubation or death at the end of the trial.

Tables with descriptive statistics of each parameter by median and interquartile range at each time point and treatment. Assessment of normality using quantile plots and log transformation to avoid lack of normality.

Kaplan-Meyer plot was used to illustrate the time to high-flow failure. The log-rank test was used to compare survival curves by a study group. The Cox regression model was used to estimate the hazard ratio for high-flow failure for sex, age, smoking status, Charlson score, diabetes, obesity, HTA, dyslipidemia, Barthel index, SOFA score, P/F, ROX, hemoglobin, hematocrit t, ferritin, leukocytes, platelets, fibrinogen, dimer, glucose, creatinine, albumin, PCR, IL-6, LDH, remdesivir, dexamethasone, PH, PCO, PO2, HCO, EB, and saturation. All were measured or collected at baseline. All analyses performed using R version 4.3.0. Main packages used were dplyr, gtsummary, ggplot2, survival, cmprsk, SPSS 25 and sjtPlot.

## RESULTS

### 394 patients were admitted

Due to exclusion criteria, 139 patients were included in the study. They had missing data, specifically 12.5% for LDH and 5% arterial blood gas (ABG) 1h after starting the treatment. They were interpolated by multiple imputations. 11 patients had DNI orders at the starting point of the study. The mean age of the population was 61.7 years, 33,8% were women. The mean P/F ratio previous starting HFT was 159. The number of comorbidities was low. The major part presented bilateral infiltrates in the X chest. The description of the population depending on the HFT outcome is in Table 1. In the cohort, 72 patients experienced HFT failure and had to switch to NPPV. In the graph (Fig. 1) we see the patients throughout the follow-up who remain on HFT. The probability of success begins to decrease from day 5 to day 10, when most HFT failures occur, being very infrequent in the following days (10 to 20). Patients who were able to withdraw treatment early due to improvement have not been excluded. 36 patients who went to trial of NPPV experienced failure and underwent mechanical ventilation. 4 patients with DNI orders died. Of the intubated patients, 6 patients died 90 days after inclusion. 10.07% was the total mortality rate at day 90 and 74% of survivors fully recovered after 1 year follow-up.

**Table 1.** Population Characteristics in subgroups, High Flow Therapy (HFT) success (no need to escalation to Non-Invasive Positive Pressure Ventilation) and HFT failure (need to escalation) Abbreviations: CRP: C-Reactive Protein; IL-6: Inteleukine-6, LDH: Lactate dehydrogenase.

|  | HFT success (n=67) | HFT failure (n=72) | p-value |
| --- | --- | --- | --- |
| Characteristics |  |  |  |
| Female | 27 (40.30%) | 20 (27.78%) | 0,12 |
| Age, Mean (SD) | 58.93 (14.82) | 64.21 (11.50) | 0,028 |
| Barthel (100p) | 60 (89.55%) | 63 (87.50%) | 0,70 |
| Smoking, n (%) | 20 (29.85%) | 27 (37.50%) | 0,30 |
| Obesity, n (%) | 23 (34.33%) | 26 (36.11%) | 0,80 |
| Hypertension, n (%) | 29 (43.28%) | 37 (51.39%) | 0,30 |
| Dislipemia, n (%) | 31 (46.27%) | 29 (40.28%) | 0,50 |
| Charlson score, Mean (SD) | 3.07 (2.51) | 4.03 (2.85) | 0.029 |
| Do-not intubate orders | 3 (4.48%) | 6 (8.33%) | 0.34 |
| Initial Clinical Status |  |  |  |
| Days from onset of symptoms, Median (IQR) | 10 (7, 12) | 8 (5.75, 10) | 0.006 |
| Sofa, Mean (SD) | 2.58 (1.49) | 3.32 (1.80) | 0.006 |
| P/F ratio, Median (IQR) | 166 (140, 200) | 140 (118, 171.5) | <0.001 |
| Chest X-ray with bilateral infiltrates, n (%) | 61 (91.04%) | 69 (95.83%) | 0.123 |
| Previous Treatment |  |  |  |
| Dexamethasone, n (%) | 67 (100.00%) | 71 (98.61%) | >0.9 |
| Enoxaparin, n (%) | 67 (100.00%) | 69 (95.83%) | 0.17 |
| Remdesivir, n (%) | 40 (59.70%) | 28 (38.89%) | 0.014 |
| Blood Test, Median (IQR) |  |  |  |
| Hemoglobin (g/dL) | 13.50 (12.60, 14.65) | 13.75 (12.50, 14.73) | >0.9 |
| Hematocrit (%) | 39.50 (37.75, 43.15) | 40.05 (36.10, 42.68) | 0.70 |
| Leukocytes (x10<sup>9</sup>/L) | 6.90 (5.50, 9.45) | 7.05 (5.15, 9.45) | 0.70 |
| Platelets (x10<sup>9</sup>/L) | 222.50 (179.00, 293.25) | 206.00 (154.00, 247.00) | 0.054 |
| Fibrinogen (mg/dL) | 700 (634.00, 803.00) | 703 (665.00, 836.75) | 0.40 |
| D-Dimer (ng/mL) | 563 (416.25, 883.75) | 598 (441.00, 930.00) | 0.40 |
| Glucose (mg/dL) | 123 (110, 156) | 147 (117.25, 197.75) | 0.022 |
| Creatinine (mg/dL) | 0.74 (0.63, 0.93) | 0.79 (0.66, 1.10) | 0.074 |
| Albumin (g/L) | 35 (32.70, 37.60) | 34.30 (32.60, 38.30) | 0.80 |
| CPR (mg/dL) | 73.30 (27.95, 125.05) | 96.70 (62.30, 138.20) | 0.085 |
| IL-6 (pg/mL) | 26.99 (8.68, 89.68) | 63.06 (41.65, 121.35) | 0.007 |
| LDH (U/L) | 322 (266.00, 392.50) | 344.50 (298.75, 454.75) | 0.055 |
| Ferritin (ng/mL) | 822 (521.50, 1,618.50) | 1,249.00 (735, 1,906) | 0.041 |

**Fig 1.**
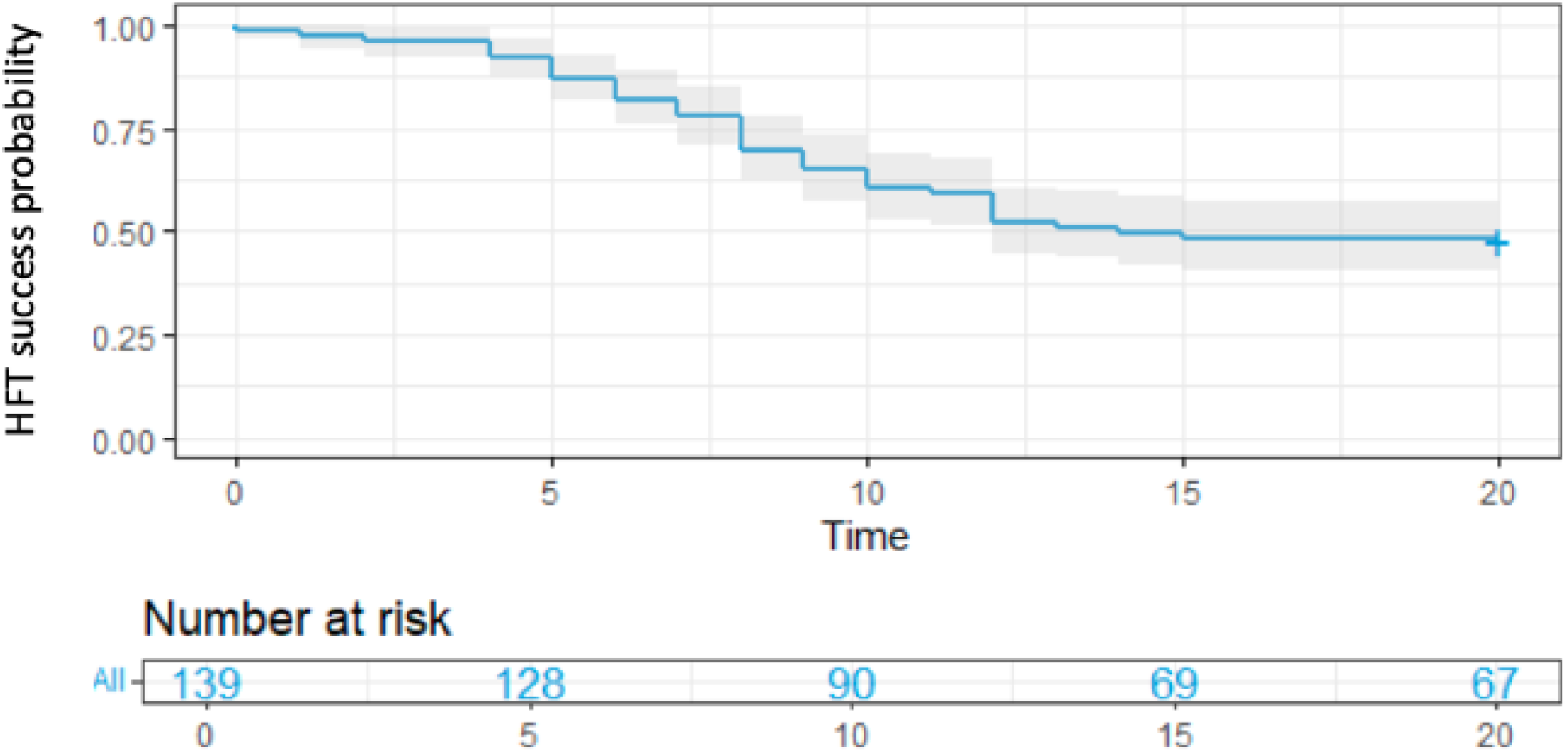
Kaplan-Meyer curve. High Flow Therapy success population who did not need escalation to Non-Invasive Positive Pressure Ventilation due to respiratory deterioration.

### Univariate Study

Cardiovascular risk factors such as smoking, obesity, hypertension, and dyslipidemia showed no significant differences between the groups. Age, Charlson score, SOFA score, P/F, had significant differences between groups. Remdesivir use was more frequent in the successful HFT group. There are differences in some blood parameters such as ferritin, Il-6 and glucose. See table 1.

### Multivariate Study

See Fig 2. A one-unit increase in the SOFA score was associated with a 1.30-fold increased risk of HFT failure (HR=1.30, 95% CI: 1.14-1.48). This finding is consistent with the qSOFA results and further emphasizes the association between organ dysfunction and HFT failure. Higher levels of LDH, CRP, Creatinine, albumin, glucose and lower levels of base excess, bicarbonate, are independent risk factors associated with HFT failure. Greater level of oxygenation or ROX index are independent protective factors with HFT failure.

**Fig 2.**
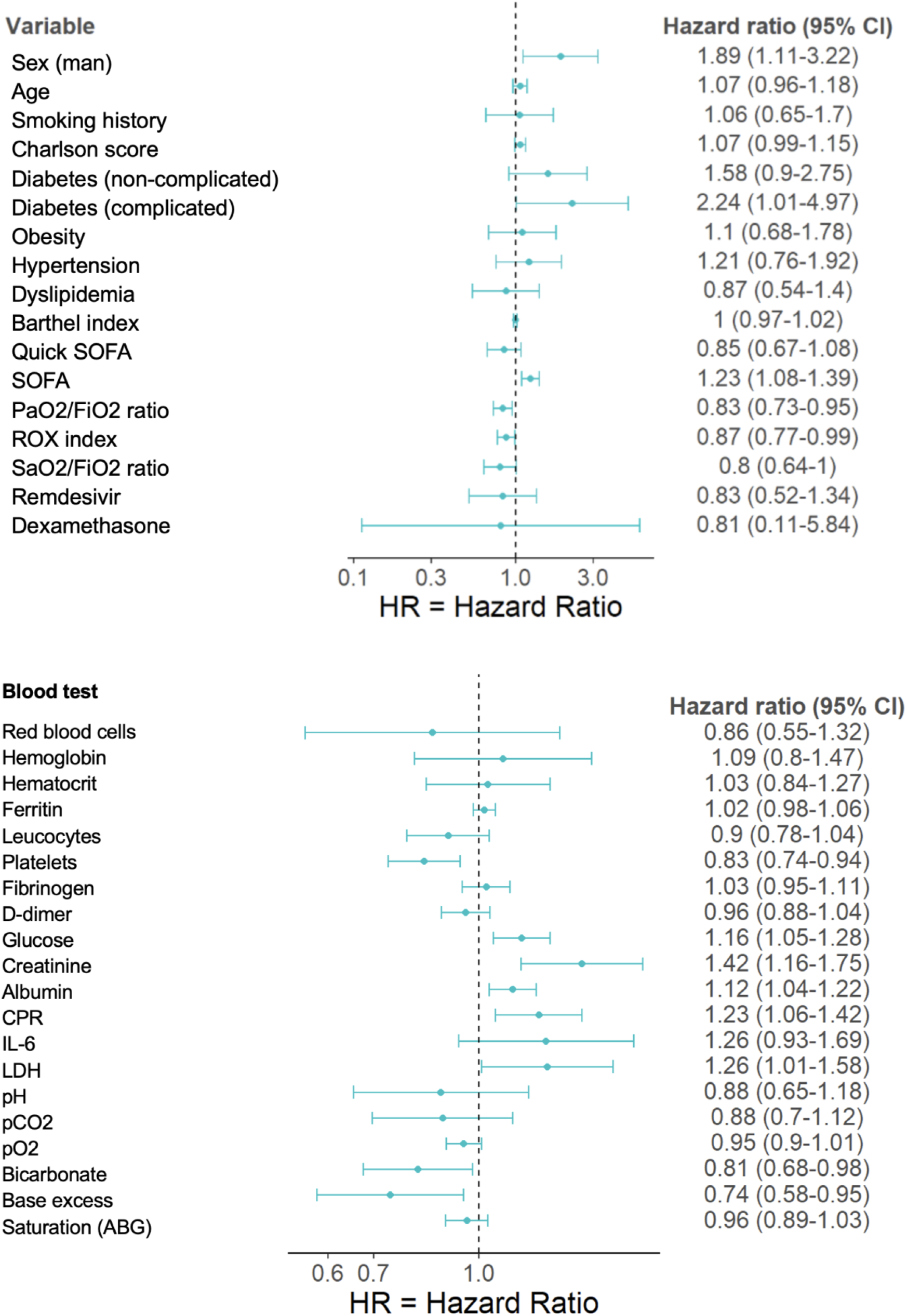
Multivariate Analysis. Risk factors associated with High Flow Therapy failure in COVID-19 Pneumonia. Hazard Ratio (95% confidence interval). Abbreviations: CRP: C-Reactive Protein; IL-6: Inteleukine-6, LDH: Lactate dehydrogenase, EB: Base excess.

## DISCUSION

The remarkable result of our study is statistically significant differences have been found in analytical biomarker values to early predict HFT failure. Regarding other severity markers described above, CRP and LDH in our prospective cohort have shown statistically significant differences when we asked about prediction of HFT failure and need to escalation to NPPV. There are expected results regarding known prognostic factors that support the previous evidence; published in retrospectives studies, (13,24) baseline non-respiratory SOFA score and ROX index were good indicators of HFT failure. (17,18).

The obtained data suggest that we must pay attention to the days from the onset of the disease because patients who started HFT treatment earlier in the course of their illness may have a lower risk of failure. We should perform close monitoring during the first 5 days after the initiation of HFT because the majority of therapy failures occur from that day. The course of the disease is a crucial point seeing that has already been demonstrated there is an increase in mortality when there is a delay in the escalation of respiratory support.

In terms of comorbidities, the better functional status is a protective against HFT failure. A one-unit increase in P/F was associated with a 0.97-fold decreased risk of HFT failure (HR=0.97, 95% CI: 0.95-0.99). A one-unit increase in ROX was associated with a 0.98-fold decreased risk of HFT failure (HR=0.98, 95% CI: 0.97-0.99). This suggests that better respiratory mechanics and oxygenation efficiency, are associated with a lower risk of HFT failure. For our sample, we could suggest a cut-off point in P/F of 150 to decide the initiation of respiratory support with NPPV.

When we evaluate blood analysis at starting HFT and after 48 hours, we find a significant decrease in D-dimer levels in the successful HFT group, suggesting reduced fibrinolysis. In the HFT failure group a significant increase in IL-6, Ferritin, and CRP were observed, indicating a more pronounced inflammatory response. A slight increase in LDH potentially could reflect tissue damage. An increase in glucose was observed in both groups, with a more pronounced rise in the HFT failure group, this could reflect stress hyperglycemia. Increases in urea and creatinine indicating potential kidney dysfunction. A significant increase in troponin suggesting potential myocardial injury. (See supplementary data)

Regarding the biomarkers, in April 2020, the first systematic reviews were already published demonstrating that along with age and comorbidities high CRP and LDH values are predictors for any outcomes for novel COVID19 pneumonia. (25) As the disease progresses, there is also a downward trend for lymphocytes, prealbumin, and albumin, whereas the opposite is seen for white blood cell count, neutrophil count, CRP, and LDH. (26) Other significant biomarkers in the previously published bibliography, include increased serum amyloid A (SAA), neutrophil-to-lymphocyte ratio (NLR), inflammatory cytokines IL-2R, tumor necrosis factor-α (TNF-α), and IL-10 (27). A limitation of our study is that these markers, being economically restricted, have not been studied. The association between serum viral load, disease severity, and outcome has also been analyzed in several studies. This factor has not been measured in our study either. The selection of patients, is very important in this kind of study. Since as we select a population with a high degree of severity, the differences in biomarkers are scarcer compared to the general population. Therefore, studies of prognostic biomarkers in specific population groups are necessary.

CRP is a sensitive inflammatory marker that increases in response to interleukin-6 (IL-6) during infectious processes. Immune and inflammatory system dysregulation is associated with disease severity. IL-6 induces CRP gene expression and the cytokine storm in COVID-19 justifies its use as a predictor of cytokine release syndrome.(28,29) It is described, a CRP level ≥75 mg/dL in Caucasic population is associated with clinical deterioration. The study of *Nazemi* establishes a cut-off point for initiating anti-inflammatory therapy. (30) Their meta-analysis showed that this increase was significantly associated with adverse clinical outcomes, including ICU admission and death. CRP levels were 0.87 times higher in patients with severe COVID-19 than in patients without severe COVID-19 and 1.32 times higher in non-survivors than in survivors. More precisely, the higher the CRP level, the worse the prognosis. In recent years, large numbers of CRP deposits have been found in inflammatory lesions of vascular endothelium infected with pathogens ARDS Clinical Practice Guideline 2021 suggest against using serum C-reactive protein and procalcitonin levels to identify bacterial pneumonia as the underlying disease. (31) Therefore, we could also infer that a component of bacterial superinfection may be added in patients who are more seriously ill.

Only 2 previous studies correlated HFT and CRP in COVID19. One small study showed that reduced serum CRP on day 4 significantly predict HFNC effectiveness in COVID-19 patient. (32) The second one, in a Japanese a retrospective cohort study published on august 2023, concluded that higher CRP level were significant factors for HFT failure and 28-day mortality. The stablished CRP level for HFT failure was > 10.0 mg/dL (cut-off value, 9.7–11 mg/dL. (33–35)

This same study shows that lactate dehydrogenase (LDH) level > 450 IU/L (cut-off value, 245–450 U/L) as the risk factors for respiratory failure or death. (33,35,36) LDH was considered as independent predictive laboratory parameters for assessing the severity of COVID-19, in which an early decline may be related to better outcomes. (37) Notably, gradually decline of LDH occured within 10 days after admission in noncritical cases but not in critical or deceased cases throughout the course of illness Since higher levels of LDH had been observed in nonsurvivors at the early stage of the illness, (38) measuring it on admission will be of greater predictive value for patients’ risk rather than during ICU.

High LDH has also been associated with worse outcomes in several studies, (14,15,39) According to a first meta-analysis (40) the mean value of LDH in severe patients was 1.54 times higher than in non-severe cases (344.48U/L vs 224.20U/L; 95%CI: 307.08−381.88U/L and 205.33−243.07U/L, respectively). Additionally, elevated baseline LDH levels were significantly associated with risk of ARDS (HR: 1.61; 95%CI: 1.44−1.79) and mortality (HR: 1.30; 95%CI: 1.11−1.52). (41) Second Meta-analysis (42) confirmed that lactate dehydrogenase level can be used as a COVID-19 severity marker and is a predictor of survival. Also between admission and emergency department regarding the need for orotracheal intubation, treatment with HFT (43) or non-invasive mechanical ventilation (44);

LDH is an intracellular enzyme present in humans in five separate isozymes (LDH-1 in cardiomyocytes, LDH-2 in reticuloendothelial system, LDH-3 in pneumocytes, LDH-4 in kidneys and pancreas, and LDH-5 in liver and striated muscle). (45) LDH abnormal values can result from multiple organ injury and decreased oxygenation with upregulation of the glycolytic pathway. The acidic extracellular pH due to increased lactate from infection and tissue injury triggers the activation of metalloproteases and enhances macrophage mediated angiogenesis (46). Severe infections may cause cytokine-mediated tissue damage and LDH release (46). Since LDH is present in lung tissue (isozyme 3), patients with severe COVID-19 infections can be expected to release greater amounts of LDH in the circulation, as a severe form of interstitial pneumonia, often evolving into acute respiratory distress syndrome, is the hallmark of the disease. Additionally, LDH levels are elevated in thrombotic microangiopathy, which is associated with renal failure and myocardial injury (47,48)

When compared to other biomarkers (low lymphocyte counts and CRP), high level of LDH was the most valuable predictive factor for mortality. (49)

Regarding the monitoring, the evolution of analytical parameters in patients undergoing HFT therapy provides valuable information for risk stratification and potential prognostication of HFT failure. The data is limited to two time points (baseline and 48 hours) and does not provide a comprehensive picture of the longitudinal changes in these parameters. The sample size might not be large enough to draw definitive conclusions about the association of these changes with HFT outcomes. Other factors, such as patient demographics, comorbidities, and medications, could also influence the observed changes in these parameters.

Larger, prospective studies with longer follow-up periods are needed to confirm these findings and identify the most predictive parameters for HFT failure. Investigating the mechanisms underlying the observed changes in these parameters could provide insights into the pathophysiology of HFT failure and potential therapeutic targets. Stratifying the analysis by patient characteristics and HFT settings could reveal more specific associations between parameter changes and outcomes

## CONCLUSION

A part from sepsis and respiratory parameters, high level of CRP and LDH, early on in the course of the disease can be a good predictor of lung injury and severe COVID-19 cases. Facilitating the selection of patients who require greater respiratory support is important to avoid an increase in mortality due to delayed intubation. For the first time we have shown a prospective study were CPR and LDH predict patients could benefit of earliest introduction of HFT with respect to adding or switching to NPPV. Specifically, when you are using a respiratory support escalation strategy. The simplicity and stability of CRP and LDH determination offer more precision medicine in clinical management compared to current predictors.

Immediate application of predictive factors would improve the clinical care of patients and allow anticipation of those who need to be transferred to a critical care unit, avoiding a delay in the onset of invasive mechanical ventilation. At the same time it avoids moving patients in whom the HFT is effective, which lowers the costs of treatment. Enhancing the figure of intermediate respiratory care units Multicentric prospective studies with longer follow-up periods are needed to confirm these findings

## Data Availability

The data that support the findings of this study are available on request from the corresponding author. The data are not publicly available due to privacy or ethical restrictions.

## AKNOWLEDGE

To all healthcare team from Hospital Germans Trias I Pujol COVID-19 Respiratory Intermediate Care Unit.

